# FootNet: A Multi-View Smartphone Dataset and Four-Model Benchmark for Clinical Foot Segmentation

**DOI:** 10.64898/2026.07.15.26358117

**Authors:** Adith Vijay, Akash Prabhune, Vinay R Srihari, Akhila Rayampalli

## Abstract

We present FootNet, a 453-image multi-view smart-phone foot dataset for binary foot segmentation, with expert-annotated masks across six anatomical views (dorsal, medial, and plantar, both left and right). We benchmark four segmentation models under a controlled protocol: U-Net with a MobileNetV2 encoder achieves the best performance (IoU 0.9268, Dice 0.9608, 95% CI [0.9209, 0.9320]); DeepLabV3 with MobileNetV3-Large scores IoU 0.8984 (Dice 0.9449); UNet++ with MobileNetV2 scores IoU 0.8913 (Dice 0.9391); and SAM ViT-B with oracle bounding-box prompt scores IoU 0.9219 on the matched 191-image subset. Bonferroni-corrected Wilcoxon signed-rank tests (*k* = 6 comparisons) show U-Net significantly outperforms DeepLab (*p*< 0.001, *r* = 0.638) and SAM ViT-B with oracle bounding-box (*p* = 0.005, *r* = 0.202); UNet++ does not significantly differ from DeepLab (*p* = 0.062). Connected-component post-processing yields negligible benefit (mean ΔIoU = +0.0003, 12 of 453 images improved). The extended dataset is available upon request.

## I. Introduction

Automated whole-foot segmentation from consumer smart-phone photographs is an enabling preprocessing step for clinical workflows including wound area measurement, deformity grading, footwear fitting, and longitudinal shape tracking. Real-world clinical photographs are captured hand-held under ambient lighting against varied, often skin-coloured backgrounds, making the task substantially harder than controlled-environment foot imaging acquired with 3D plantar scanners [1], thermal cameras [2], or depth sensors.

Existing public foot datasets target wound-level segmentation rather than the whole foot. The Diabetic Foot Ulcer Challenge (DFUC) [3], [4] is the primary benchmark for ulcer-region masks; prior deep-learning surveys cover wound detection and healing prediction from smartphone images [5], [6], but whole-foot segmentation masks are not released. To our knowledge, no publicly available multi-view whole-foot dataset acquired under realistic point-of-care conditions existed prior to our initial release.

This paper makes four contributions: (1) FootNet, a 453-image multi-view smartphone foot dataset with expert-annotated binary masks (191 images publicly available at DOI: 10.5281/zenodo.20457252; 262 images available upon request pending de-identification review); (2) a controlled four-model benchmark comparing U-Net [7], UNet++ [8], DeepLabV3-MobileNetV3 [9], [10], and SAM ViT-B [11]; (3) Bonferroni-corrected Wilcoxon signed-rank significance tests across all six model pairs with bootstrap 95 % confidence intervals and rank-biserial effect sizes; and (4) a connected-component post-processing ablation.

## II. Related Work

### A. Foot Datasets and Segmentation

Smartphone-based diabetic foot ulcer analysis has been explored in [3]–[6], establishing consumer cameras as a clinically viable modality. However, these works target the wound region and not the whole foot. Hardware-dependent approaches for whole-foot analysis — 3D plantar scanners [1] and thermal cameras [2] — achieve sub-millimetre accuracy but require dedicated equipment unavailable at the point of care.

### B. Segmentation Architectures

U-Net [7] introduced encoder-decoder skip connections that have become the dominant paradigm for medical image segmentation. UNet++ [8] extends U-Net with nested dense skip pathways to reduce the semantic gap between encoder and decoder levels. DeepLabV3 [9] uses atrous spatial pyramid pooling for multi-scale context and, paired with MobileNetV3-Large [10], reaches near-ResNet50 segmentation accuracy at approximately 11 million parameters. The Segment Anything Model (SAM) [11] is a prompt-based vision foundation model trained on over one billion masks; its zero-shot performance on medical images is uneven [12], [13], with domain adaptation often required for reliable results [14], [15].

## III. Dataset: Footnet

### A. Acquisition and Annotation

Images were collected at outpatient clinics by clinic staff using various consumer smartphones, with no tripods, controlled lighting, or standardised backdrops. Resolutions range from 1080 px to 4160 px on the longer axis. Images were annotated by trained annotators using LabelMe, tracing a polygon along the entire foot boundary; LabelMe JSON annotations were converted to binary PNG masks using OpenCV’s `fillPoly` (foreground = 255, background = 0). JPEG images were loaded with PIL’s `ImageOps.exif_transpose` to apply EXIF rotation metadata before annotation alignment. EXIF metadata and any personally identifiable information visible in frame were removed before release.

### B. Composition

FootNet comprises 453 image–mask pairs across twelve view folders corresponding to six anatomical views (Table I). The original 191-image set (Footnet Dataset; folder names *dataset left/righttop/side/sole*) was extended by 262 new images (Footnet Dataset new; folder names *Left/Rightdorsal, Left/Rightmedial, Left/Rightplantar*), where top = dorsal, side = medial, and sole = plantar.

**TABLE I.** FootNet Dataset Composition.

| Anatomical view | FD | FDn | Total |
| --- | --- | --- | --- |
| Left dorsal | 30 | 44 | 74 |
| Left medial | 31 | 43 | 74 |
| Left plantar | 33 | 44 | 77 |
| Right dorsal | 39 | 44 | 83 |
| Right medial | 28 | 44 | 72 |
| Right plantar | 30 | 43 | 73 |
| **Total** | **191** | **262** | **453** |

FD=Footnet\_Dataset; FDn=Footnet\_Dataset\_new.

### C. Data Splits and Availability

For supervised model training, samples are split per view folder with `val_ratio = 0.15` and `seed = 42` (stratified random split), yielding reproducible train and validation sets. SAM is evaluated on the original 191 images; no training is required for that model. The original 191-image Foot-net Dataset is publicly available at https://doi.org/10.5281/zenodo.20457252 (CC BY 4.0). The extended 262-image Footnet Dataset new is undergoing de-identification review prior to public release; it is available to researchers upon request to the corresponding author.

## IV. Methods

### A. U-Net and UNet++

We use the `segmentation-models-pytorch` library with a MobileNetV2 encoder pre-trained on ImageNet for both U-Net [7] and UNet++ [8]. All images are resized to 384 × 384 (stride-32 aligned for MobileNetV2’s downsampling factor). Training uses a combined Dice + BCE loss (0.5 : 0.5), AdamW (lr = 10^−3^, weight decay 10^−4^), cosine-annealing schedule (*T*_max_ = 60, *η*_min_ = 10^−6^), batch size 4, mixed-precision (FP16 autocast with gradient scaling), and early stopping with patience 12 on validation Dice.

### B. DeepLabV3-MobileNetV3-Large

We fine-tune `torchvision`’s `deeplabv3_mobilenet_v3_large` (COCO pre-trained, approximately 11 million parameters, 45 MB) by replacing both the 21-class main classifier head and auxiliary head with `Conv2d(·, 1, kernel_size=1)` layers for single-channel binary output. The auxiliary head contributes 0.4 × the main Dice+BCE loss during training only. All other training hyperparameters match Section IV-A.

### C. SAM ViT-B Baseline

SAM ViT-B (90 million parameters, 375 MB) is evaluated zero-shot using the official checkpoint [11] and one oracle bounding-box prompt per image: the tight bounding box of the ground-truth mask padded 20 px on each side. This protocol is an explicit upper bound on prompt quality; real-world SAM IoU under a predicted box will be lower. SAM is evaluated on the original 191 images only.

### D. Augmentation and Evaluation Protocol

Training augmentations (Albumentations): horizontal flip (*p* = 0.5), vertical flip (*p* = 0.2), RandomRotate90 (*p* = 0.3), affine perturbation (translate ±8 %, scale 0.85–1.15, rotate ± 20°, *p* = 0.6), one of RandomBrightnessContrast or Hue-SaturationValue (*p* = 0.5), Gaussian noise (*p* = 0.2), and ImageNet normalisation. Validation uses resize + normalize only.

Per-image IoU and Dice are computed at the original image resolution. Wilcoxon signed-rank tests compare matched image-level IoU scores; all reported *p*-values are two-sided. Six pairwise comparisons are performed (three among supervised models on *n*=453, three involving SAM on the *n*=191 subset); Bonferroni correction sets the adjusted significance threshold at *α*_Bonf_ = 0.05*/*6 = 0.0083. Effect size is reported as rank-biserial correlation 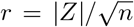, where *Z* is the standard-normal approximation to the Wilcoxon statistic. Bootstrap 95 % confidence intervals use 10 000 resamples (`seed = 42`, percentile method).

## V. Results

### A. Model Comparison

Table II reports mean IoU and Dice with 95 % bootstrap confidence intervals. The three trained models are evaluated on all 453 images; SAM is evaluated on the matched 191-image subset only. Fig. 1 summarises per-model mean IoU with confidence intervals and pairwise significance.

**TABLE II.** Segmentation Performance on FootNet. IoUa : FULL 453-IMAGE SET (*n*=453); IoU_B_: MATCHED 191-IMAGE SAM SUBSET (*n*=191).

| Model | Dice | IoU<sub>A</sub> | 95 % CI | IoU<sub>B</sub> |
| --- | --- | --- | --- | --- |
| U-Net (MBV2)* | **0.9608** | **0.9268** | [0.9209, 0.9320] | 0.9274 |
| DeepLab (MBV3)* | 0.9449 | 0.8984 | [0.8920, 0.9040] | 0.9090 |
| UNet++ (MBV2)* | 0.9391 | 0.8913 | [0.8818, 0.8999] | 0.9058 |
| SAM ViT-B† | — | — | [0.9070, 0.9340] | 0.9219 |

\*Supervised; trained on FootNet ( $n=453$ ).
†Zero-shot; oracle bounding-box prompt ( $n=191$ ).
MBV2 = MobileNetV2; MBV3 = MobileNetV3-Large.

**Fig. 1.**
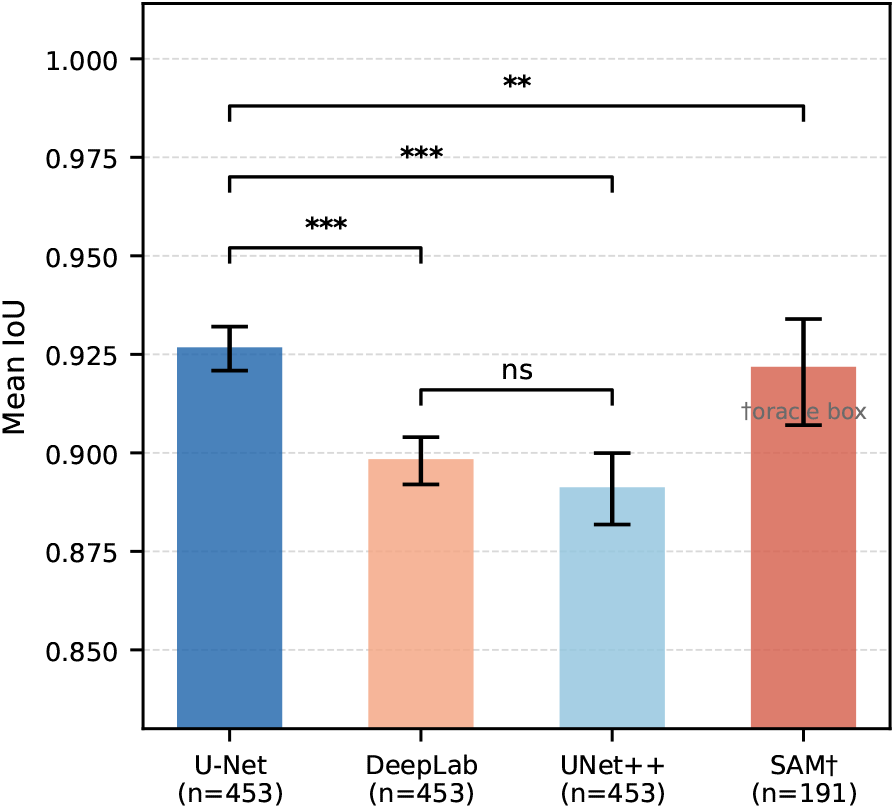
Mean IoU with 95 % bootstrap confidence intervals for each model. Significance brackets show Wilcoxon signed-rank results (∗ ∗ ∗ *p*<0.001; ∗∗ *p*<0.01; ns = not significant, *p*=0.062). SAM († ) is evaluated on the matched 191-image oracle-box subset only.

U-Net achieves the highest IoU (0.9268) and Dice (0.9608). Bonferroni-corrected Wilcoxon signed-rank tests (*α*_Bonf_ = 0.0083, *k* = 6 comparisons) on per-image IoU show large effects for both trained-model comparisons: U-Net outperforms DeepLab (*p*< 0.001, *r* = 0.638) and UNet++ (*p*< 0.001, *r* = 0.646). DeepLab versus UNet++ is not significant after correction (*p* = 0.062, *r* = 0.088). On the matched 191-image SAM subset (IoU_B_, Table II), U-Net (0.9274) significantly outperforms SAM (0.9219, *p* = 0.005, *r* = 0.202); DeepLab (0.9090) is significantly outperformed by SAM (*p*< 0.001, *r* = 0.354); and the UNet++ (0.9058) versus SAM comparison does not survive Bonferroni correction (*p* = 0.036 > 0.0083, *r* = 0.151).

### B. Connected-Component Ablation

Retaining only the largest 8-connected component of each DeepLab prediction produced a mean IoU change of +0.0003 across all 453 images, with only 12 of 453 images showing any improvement. CCA is therefore not adopted as a default post-processing step.

### C. Per-View Breakdown

Table III shows U-Net IoU aggregated by anatomical view. Plantar views achieve the highest accuracy (left plantar 0.9526, right plantar 0.9496); dorsal and medial views are 2–4 % lower. Right dorsal has notably higher variance (std 0.102), driven by a subset of images with complex backgrounds. The qualitative panel in Fig. 2 illustrates four cases spanning the range of model agreement: a best-case plantar view where all models agree (IoU 0.97–0.98), a typical medial view, a challenging left-dorsal scene where U-Net (IoU 0.868) is substantially more robust than DeepLab (0.655) and UNet++ (0.401), and a right-dorsal case where DeepLab (0.938) and UNet++ (0.941) outperform U-Net (0.876), demonstrating that U-Net’s aggregate superiority does not hold on every image.

**TABLE III.** Per-View U-Net IoU (*n* = 453)

| View | n | Mean IoU | Std |
| --- | --- | --- | --- |
| Left plantar | 77 | 0.9526 | 0.029 |
| Right plantar | 73 | 0.9496 | 0.038 |
| Right medial | 72 | 0.9230 | 0.039 |
| Left dorsal | 74 | 0.9214 | 0.052 |
| Left medial | 74 | 0.9210 | 0.045 |
| Right dorsal | 83 | 0.8960 | 0.102 |

**Fig. 2.**
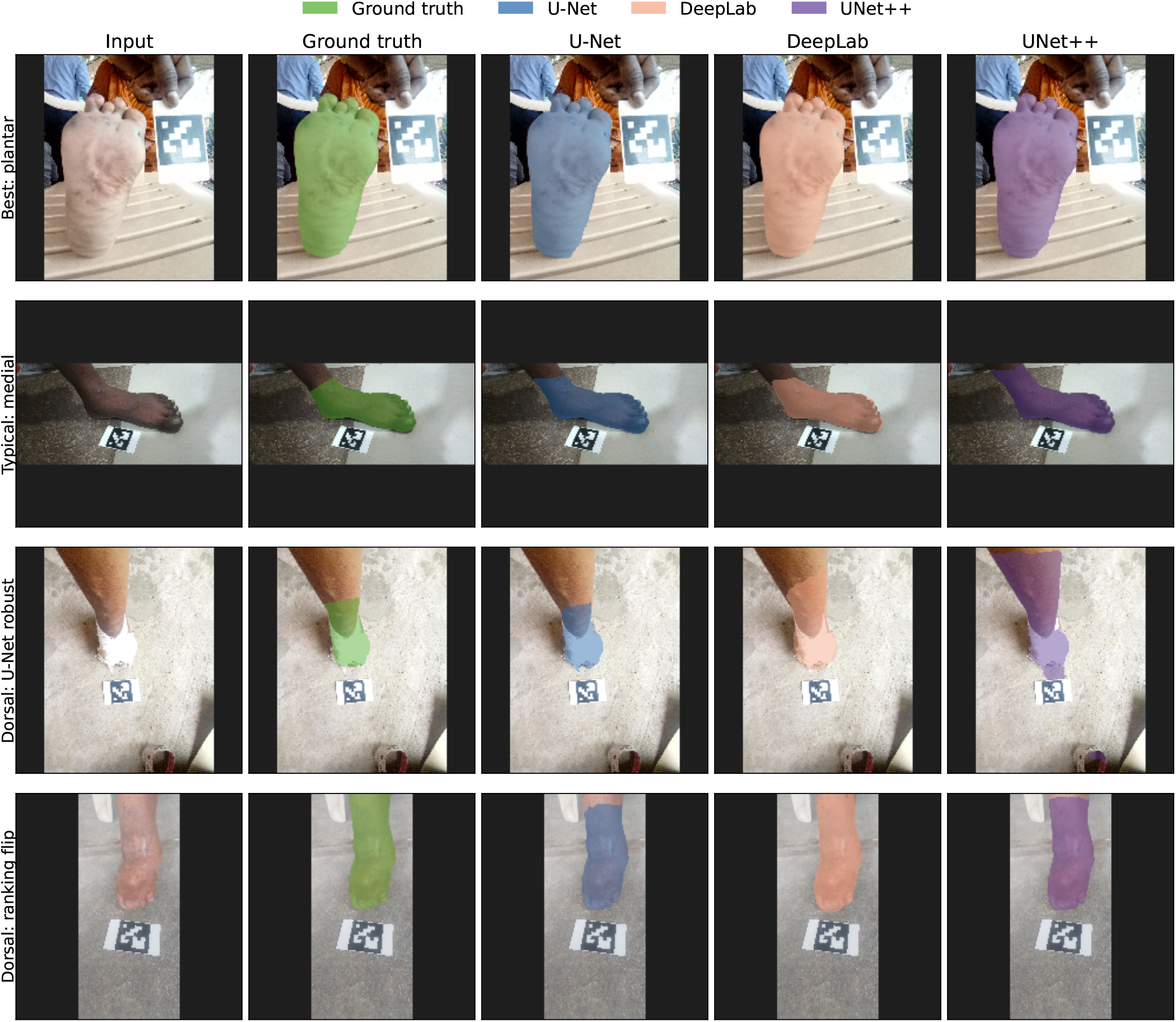
Qualitative segmentation comparison (4 rows× 5 columns). Columns: original image, ground-truth mask (green), U-Net (blue), DeepLab (salmon), UNet++ (purple). **Row 1 (best case):** right-plantar view; all three models agree (U-Net 0.982, DeepLab 0.974, UNet++ 0.981). **Row 2 (typical):** left-medial view near the dataset median; U-Net and DeepLab agree (IoU 0.911) while UNet++ scores lower (0.827). **Row 3 (dorsal — U-Net robust):** left-dorsal view with cluttered background; U-Net IoU 0.868 versus DeepLab 0.655 and UNet++ 0.401. **Row 4 (dorsal — ranking flip):** right-dorsal view where DeepLab (0.938) and UNet++ (0.941) outperform U-Net (0.876), illustrating that aggregate superiority does not hold on every image.

## VI. Discussion

U-Net with a MobileNetV2 encoder is the strongest model across all comparisons, outperforming both DeepLabV3 and SAM with oracle prompts. This aligns with U-Net’s established dominance in medical image segmentation [7]: the encoder-decoder skip connections preserve fine boundary detail that atrous pooling at 384 × 384 resolution partially discards. The finding that UNet++ does not significantly out-perform DeepLab (*p* = 0.062) — while U-Net significantly outperforms UNet++ — suggests that the nested dense skip pathways add complexity without a measurable accuracy benefit at this dataset scale and resolution.

SAM with oracle bounding-box prompts (IoU_B_ 0.9219) outperforms DeepLab (0.9090, *p*< 0.001) on the matched 191-image subset but is significantly outperformed by U-Net (0.9274, *p* = 0.005, *r* = 0.202, both Bonferroni-corrected). The UNet++ versus SAM difference (*p* = 0.036) does not survive Bonferroni correction and should not be interpreted as a reliable performance gap at this sample size. SAM’s higher variance reflects its sensitivity to within-box ambiguity — a property not fully mitigated even when the box is derived from the ground-truth mask. For downstream pipelines, U-Net offers both higher mean accuracy and lower worst-case failure risk, and operates prompt-free at a fraction of SAM’s 375 MB checkpoint size.

Plantar views consistently achieve the highest segmentation accuracy (left plantar 0.9526, right plantar 0.9496) across all models, likely because the foot sole presents a convex, high-contrast silhouette against a floor or table. Right-dorsal images show elevated variance (std 0.102), which we attribute to greater background complexity and a wider range of patient postures in that view.

## Data Availability

The original 191-image FootNet dataset (Footnet_Dataset) is publicly available at Zenodo (DOI: 10.5281/zenodo.20457252, CC BY 4.0). The extended 262-image subset (Footnet_Dataset_new) is currently undergoing de-identification review prior to public release and is available to researchers upon reasonable request to the corresponding author (Adith Vijay, ADMIRE, IIHMR Bangalore).

https://zenodo.org/records/20457253

## Limitations

The dataset has several constraints that qualify the scope of these findings. *Single site and annotators*. All 453 images originate from a single clinical site and were annotated by trained annotators. Inter-rater reliability is not reported, so annotation variability is unknown; polygon-based LabelMe tracing may introduce systematic smoothing over deformed or splayed toes. *Image-level rather than patient-level split*. Train/validation splits are performed independently within each view folder (15 % held out per folder, `seed = 42`). Because multiple images of the same patient may appear across view folders, the same patient’s foot could contribute to both training and validation. The degree of resulting data leakage is unknown without structured patient identifiers; reported validation IoU scores may therefore be modestly optimistic for deployments on wholly unseen patients. *Inconsistent naming convention*. The original and extended subsets use different view-name conventions (*top/side/sole* vs. *dorsal/medial/plantar*), which must be reconciled by any code that processes the full corpus. *Protocol mismatch between supervised models and SAM*. SAM is evaluated on the original 191-image subset only (no training required), while supervised models are evaluated on all 453 images. To mitigate this, Table II reports IoU for all three supervised models on the matched 191-image subset (IoU_B_), enabling a direct same-*n* comparison; trends are consistent with the full-set results, and Bonferroni-corrected statistics are reported throughout. *No cross-site validation*. All training, validation, and testing images come from the same site and acquisition conditions. Performance on images from different sites, lighting conditions, or devices is untested. *No on-device benchmark*. Inference times were measured on an NVIDIA RTX 4050 desktop GPU. On-device performance on representative Android or iOS hardware — the intended deployment environment — has not been measured; ONNX or TFLite export and mobile profiling are left as future work. *Limited dataset size*. With 453 images from a single site, the dataset is modest by deep-learning standards. Larger and more diverse collections would be needed to establish generalisability and to train models such as transformer-based architectures that typically require more data. *Standard architectures only*. This benchmark evaluates CNN-based encoders (MobileNetV2, MobileNetV3-Large) and the prompt-based SAM ViT-B; recent transformer-based segmentation models (e.g., SegFormer, Swin-UNet) are not included. Their performance on FootNet remains an open question. *Decision-threshold sensitivity*. All models use a fixed probability threshold of 0.5. Per-view or dataset-level threshold tuning on a held-out set could improve boundary precision, particularly for medial and dorsal views.

## VII. Conclusion

FootNet provides 453 expert-annotated smartphone foot images across six anatomical views. A controlled four-model benchmark with Bonferroni-corrected Wilcoxon tests (*k* = 6) shows that U-Net with a MobileNetV2 encoder (IoU 0.9268, Dice 0.9608) significantly outperforms DeepLabV3-MobileNetV3 (*p*< 0.001, *r* = 0.638) and SAM ViT-B with oracle prompts on the matched subset (*p* = 0.005, *r* = 0.202). UNet++ does not significantly differ from DeepLab (*p* = 0.062), and the UNet++ versus SAM comparison does not survive Bonferroni correction (*p* = 0.036). Connected-component post-processing yields negligible benefit. The original 191-image dataset is publicly available at DOI: 10.5281/zenodo.20457252; the extended set is undergoing de-identification review and is available upon request.

## Acknowledgment

The authors thank the clinicians and field staff of Lepra Society India who collected the images. We thank Hemanth B for the image annotation.

